# Droplet Digital RT-PCR to detect SARS-CoV-2 variants of concern in wastewater

**DOI:** 10.1101/2021.03.25.21254324

**Authors:** Leo Heijnen, Goffe Elsinga, Miranda de Graaf, Richard Molenkamp, Marion P.G. Koopmans, Gertjan Medema

**Affiliations:** KWR Water Research Institute, Nieuwegein, The Netherlands; Department of Viroscience, Erasmus University Medical Center, Rotterdam, The Netherlands

## Abstract

Wastewater surveillance has shown to be a valuable and efficient tool to obtain information about the trends of COVID-19 in the community. Since the recent emergence of new variants, associated with increased transmissibility and/or antibody escape (variants of concern), there is an urgent need for methods that enable specific and timely detection and quantification of the occurrence of these variants in the community. In this study we demonstrate the use of RT-ddPCR on wastewater samples for specific detection of mutation N501Y. This assay enabled simultaneous enumeration of the concentration of variants with the 501Y mutation and Wild Type (WT, containing 501N) SARS-CoV-2 RNA. Detection of N501Y was possible in samples with mixtures of WT with low proportions of lineage B.1.351 (0.5%). The method could accurately determine the proportion of N501Y and WT in mixtures of SARS-CoV-2 RNA. The application to raw sewage samples from the cities of Amsterdam and Utrecht demonstrated that this method can be applied to determine the concentrations and the proportions of WT and N501Y containing SARS-CoV-2 RNA in wastewater samples. The emergence of N501Y in Amsterdam and Utrecht wastewater aligned with the emergence of B.1.1.7 as causative agent of COVID-19 in the Netherlands, indicating that RT-ddPCR of wastewater samples can be used to monitor the emergence of the N501Y mutation in the community. It also indicates that RT-ddPCR could be used for sensitive and accurate monitoring of current (like K417N, E484K) or future mutations present in SARS-CoV-2 variants of concern. Monitoring emergence of these mutations in the community via wastewater is rapid, efficient and valuable in supporting public health decision-making.

## Introduction

The first reports of diarrhea caused by SARS-CoV-2 in COVID-19 patients and the detection of SARS-CoV-2 RNA in the feces of 30-60% of infected individuals (Jones, Baluja et al. 2020, Wang, Hu et al. 2020, Wölfel, Corman et al. 2020, Xiao, Tang et al. 2020), sparked the idea that wastewater could be used to monitor the circulation of SARS-CoV-2 at community level. Methods for the detection of SARS-CoV-2 RNA in wastewater were developed and deployed already in the early stage of the pandemic (Lodder and de Roda Husman 2020, Medema, Heijnen et al. 2020). The concentration of SARS-CoV-2 RNA in wastewater has shown to reflect and even precede the trends of the newly reported cases or COVID-19 hospitalizations (Medema, Been et al. 2020, Prado, Fumian et al. 2021). The possibility to obtain objective information about virus circulation in a community via wastewater sampling, independent of diagnostic testing availability, willingness and awareness (i.e. asymptomatic virus carriers), is currently being used to support public health decisions. In situations with low SARS-CoV-2 circulation in the community, wastewater surveillance is being used as an early warning system (Betancourt, Schmitz et al. 2020, Medema, Heijnen et al. 2020, Ahmed, Tscharke et al. 2021) enabling rapid and targeted measures to limit viral spread. The added value of wastewater surveillance has led to worldwide implementation of methods for SARS-CoV-2 monitoring in sewage (Bivins, North et al. 2020).

The occurrence of new SARS-CoV-2 variants that have unusual numbers of mutations and are associated with increased transmissibility, antibody escape or both (“Variants of Concern”, VoC), has recently gained attention (Altmann, Boyton et al. 2021, Challen, Brooks-Pollock et al. 2021, Eurosurveillance-editorial-team 2021). Three VoC lineages are currently spreading worldwide and additional variants are under consideration: Lineage B.1.1.7 was recognized first through the large scale genomics initiative in the UK, lineage B.1.351 was first recognized in South Africa and lineage P.1 emerged in late 2020 in Manaus (Brazil). These VoC’s appear to spread at higher efficiency (Davies, Abbott et al. 2021, Faria, Mellan et al. 2021, Volz, Mishra et al. 2021). In addition, lineages B.1.351 and P.1 appear to be partially resistant to neutralizing antibodies from infection, vaccination, or treatment (Garcia-Beltran, Lam et al. 2021, Wang, Liu et al. 2021). Therefore, it is of great importance to monitor the spread of these VoC in the community to understand the dynamics of transmission of VoC and be able to take appropriate public health protection measures. Currently, VoC surveillance is mainly done through whole genome sequencing of patient samples. This approach is relatively expensive, labour intensive and time consuming and may not be achievable in many parts of the world. Variant surveillance via wastewater could be a more efficient and rapid method to monitor the emergence and spread of VoC in a community. However, there are currently no methods available to quantify the presence of different VoC’s in sewage. Sequence analysis of RNA isolated from sewage can specifically detect and identify SARS-CoV-2 RNA sequence variation (Izquierdo-Lara, Elsinga et al. 2020) and sequence analysis has the potential to identify VoC in sewage. However, it is expected that this needs deep sequencing efforts and bioinformatics to find rare mutations in sewage samples which likely contain mixtures of SARS-CoV-2 lineages. The limited sequence differences between VoC and the likely presence of mixtures of sequence variants in sewage makes it difficult to design VoC specific RT-qPCR assays for sewage. In droplet digital RT-PCR (RT-ddPCR) the sample and reagents are partitioned in a large number (~10.000 to 20.000) “water in oil” droplets and PCR-reactions are performed on single molecules in individual closed droplets. This makes it possible to detect rare mutations and to discriminate closely related sequences on the base of probe binding kinetics (Pekin, Skhiri et al. 2011). This makes RT-ddPCR potentially attractive for specific detection presence and proportion of mutations that are specific to VoC’s in wastewater. In this study we evaluated the use of RT-ddPCR to detect the N501Y mutation and to determine the proportion and concentration of VoC containing N501Y in wastewater. The N501Y mutation is present in lineages B.1.1.7, B.1.351 and P.1 and leads to an amino acid change at position 501 in the receptor binding domain of the spike protein. The N501Y mutation appears to influence binding of the spike protein to the human ACE2 receptor (Liu, Zhang et al. 2021, Rynkiewicz, Babbitt et al. 2021). Validation was performed with RNA isolated from wild type virus and a B.1.351 stock and the applicability of the RT-ddPCR was studied on domestic wastewater samples from the Dutch cities of Amsterdam and Utrecht.

## Methods

### Wastewater samples

Composite, flow-dependent 24h samples were collected by the operators of the wastewater treatment plants (WWTP) that serve the cities of Amsterdam and Utrecht in the Netherlands. Samples were stored at 4 °C during 24h periods of sampling as previously described (Medema et al 2020). Samples were taken weekly from March 2020 to March 1 2021. RT-qPCR was applied on all samples, RT-ddPCR analysis was applied on the samples from the period November 9 2020 to March 1 2021.

### SARS-CoV-2 viral RNA

Reference genomic RNA from WT SARS-CoV-2 virus (WIV04/2019) and variants B.1.351 and B.1.1.7 containing the N501Y mutation was isolated from cell-cultured virus at passage 3 after inoculation of primary patient sample. RNA isolation from 0.2 TCID50 cell-cultured virus was performed by using the MagNA pure 96 (Roche diagnostics) and the total nucleic acid isolation kit (Roche Diagnostics). The sequence of the cell-cultured RNA was confirmed by whole-genome sequencing.

### Virus concentration and nucleic acid extraction

Samples were transported to the laboratory and processed as previously described (Medema et al 2020). In short, centrifugation was used to pellet larger particles, viral particles were concentrated from 50 ml supernatant by ultrafiltration through Centricon® Plus-70 centrifugal ultrafilters with a cut-off of 30 or 100 kDa, dependent on supplies (Millipore, Amsterdam, The Netherlands). Approximately 2×10^4^ genomic RNA copies from the murine coronavirus Mouse Hepatitis Virus (MHV)-A59 (obtained from Leiden University Medical Center, The Netherlands) was spiked to each concentrate and co-isolated during the extraction procedure to monitor the possible presence of RT-PCR inhibitors and measure the recovery efficiency of the extraction procedure. Nucleic acid was extracted from the concentrate using the magnetic extraction reagents of the Biomerieux Nuclisens kit (Biomerieux, Amersfoort, the Netherlands) in combination with the semi-automated KingFisher mL (Thermo Scientific, Bleiswijk, The Netherlands) as previously described. Extracted nucleic acid was eluted in a volume of 100 µl.

### RT-qPCR for Coronavirus RNA-quantification in wastewater

The N2 assay targeting a fragment of the nucleocapsid gene, as published by US CDC (US-CDC 2020), was used to quantify SARS-CoV-2 RNA in the sewage samples. Reagents and reaction conditions were as previously described (Medema et al. 2020). All RT-PCR’s were run as technical duplicates on 5 µl extracted nucleic acid. RT-qPCR reactions on serial dilutions containing RT-ddPCR calibrated EURM-019 single stranded RNA (provided by the Joint Research Centre) were used to construct calibration curves that subsequently were used to quantify N2 in RNA extracted from the sewage samples. Reactions were considered positive if the cycle threshold was below 40 cycles. Spiked MHV-A59 RNA was detected by performing a MHV-A59 specific RT-qPCR targeting the N-gene using the primers and reaction profile described by Raaben et al. (Raaben, Einerhand et al. 2007). Calibration curves for quantification were generated by performing RT-PCR assays on dilution series of a synthetic quantified gBlock gene fragment containing a partial sequence MHV-A59 N-gene (obtained from IDT, Leuven Belgium). The recovery efficiency was determined by comparing the MHV-A59 RNA concentration in the sewage sample with the concentration MHV-A59 suspension used to spike the samples. This recovery efficiency was used to monitor the performance of the RNA-isolation procedure and the possible presence of RT-PCR inhibitors. The recovery efficiencies are used as process control, but not used to correct the measured SARS-CoV-2 RNA concentrations (Kantor, Nelson et al. 2021).

### PCR for CrAssphage quantification in wastewater

A previously described CrAssphage CPQ_064 specific PCR (Stachler, Kelty et al. 2017) was used to quantify this DNA-virus that is ubiquitously present in human intestinal tracts in high concentrations. Assays were performed in duplicate on 5 µl 1:10 diluted extracted nucleic acid. Quantification was performed using PCR assays on dilution series of a synthetic quantified gBlock (obtained from IDT, Leuven, Belgium) containing the CPQ_064 gene fragment. In each wastewater sample, the measured SARS-CoV2 N2-gene concentration was divided by the CrAssphage concentration to normalize for the dilution of the human faecal load by fluctuating human and non-human inputs in the sewer network, such as rain- or groundwater (Medema, Been et al. 2020).

### RT-ddPCR for the N501Y mutation

Digital droplet RT-PCR was used to quantify the N501Y and the wild-type (*WIV04/2019*, WT) sequence in one single tube multiplex mutation assay designed by BioRad (Assay ID: dMDS731762551). This assay uses primers that amplify a 80 bp fragment of the Spike gene including the area containing an A to T point mutation that leads to the N501Y amino acid change in the Spike protein (N501Y). Two probes are used to detect PCR-amplification in the droplets: one FAM-labeled probe which perfectly binds to the N501Y mutation and one HEX-labeled probe which perfectly binds to the wild-type SARS-CoV-2 sequence. The ability to perform the PCR-assay in discrete self-contained droplets makes it possible to discriminate between droplets containing SARS-CoV-2 mutant fragments at low frequencies in a background of wild-type fragments. Assays were performed in 20 µl reaction volumes containing the reagents from the One-Step Advance RT-ddPCR for probes: 5 µl RT-ddPCR One-Step Advanced Supermix, 2 µl Reverse Transcriptase, 1 µl DTT (300 mM) supplemented with 1 µl Single tube mutation assay, 6 µl PCR grade and RNAse free water (Applied Biosystems, Fisher Scientific, Landsmeer, The Netherlands) and 5 µl sample-RNA. The BioRad QX200 droplet generator partitioned sample-RNA and reagents in droplets. The temperature profile used for RT-ddPCR was as follows: 60 min. 50°C, 10 min 50°C, 40 cycles with 30 sec. 95°C and 1 min. 55°C followed by 10 min. 98°C, 30 min. 4°C and hold at 12°C. Samples were scanned using the QX200 system (BioRad) and analyzed using the QuantaSoft-Analysis software (BioRad). For each sample, the number of negative and WT or N501Y ddPCR positive droplets were recorded and used to determine the WT or N501Y concentrations. The proportion of Spike gene specific RNA fragments containing the N501Y mutation was calculated by the QuantaSoft-Analysis software as the concentration N501Y in the ddPCR reaction, divided by the sum of WT and N501Y concentrations in the ddPCR reaction. The 95% confidence intervals in the proportion of N501Y were calculated assuming a Poisson distribution of RNA molecules in the droplets.

### Validation experiments

Two dilution series were analyzed to evaluate the ability of RT-ddPCR to detect WT and the N501Y variants simultaneously in samples with different concentration ratios of SARS-CoV-2 lineage B.1.351 and WT. The approximate concentration of RNA from WT SARS-CoV-2 virus (Wuhan type) and variant B.1.351 was first quantified using the N2 specific RT-qPCR assay. The first dilution series consisted of a stable concentration of approximately 600 RNA copies of WT, mixed with 2-fold dilutions of lineage B.1.351. The second dilution series contained a stable concentration of approximately 700 RNA copies of variant B.1.351 mixed with 2-fold dilution series of RNA extracted from WT virus. The average concentrations measured with RT-ddPCR in the samples containing stable concentrations of WT or variant B.1.351 respectively were used as values, the dilution factors were subsequently used to calculate the expected proportions of WT and variant B.1.351 in the mixed samples.

## Results

### Method validation

To study the ability of RT-ddPCR to differentiate between 501Y and 501N sequences, and to detect low concentrations of SARS-CoV-2 N501Y mutant in the background of WT RNA, two dilutions series were analyzed. For this, we used RNA isolated from cell cultures infected with wild type virus and a B.1.351 strain containing the N501Y mutation. A two-fold dilution series of B.1.351 RNA was made in a stable background of WT RNA (Figure 1) and vice versa (Figure 2), and the concentrations of WT and N501Y was determined by RT-ddPCR.

**Figure 1.**
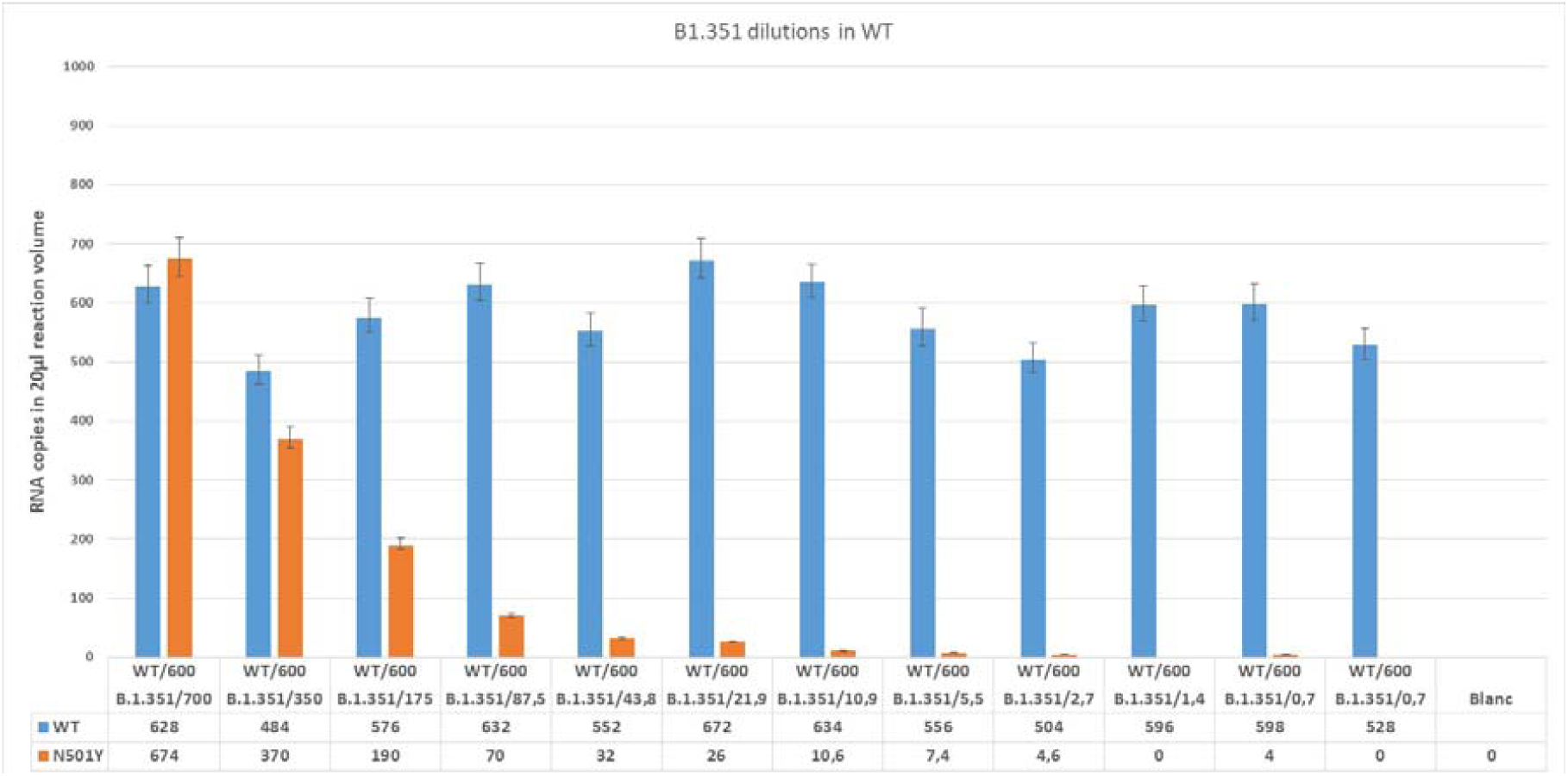
Detected copy numbers of WT and N501Y in samples containing ~600 copies WT and 2-fold dilution series of variant lineage B.1.351.

**Figure 2.**
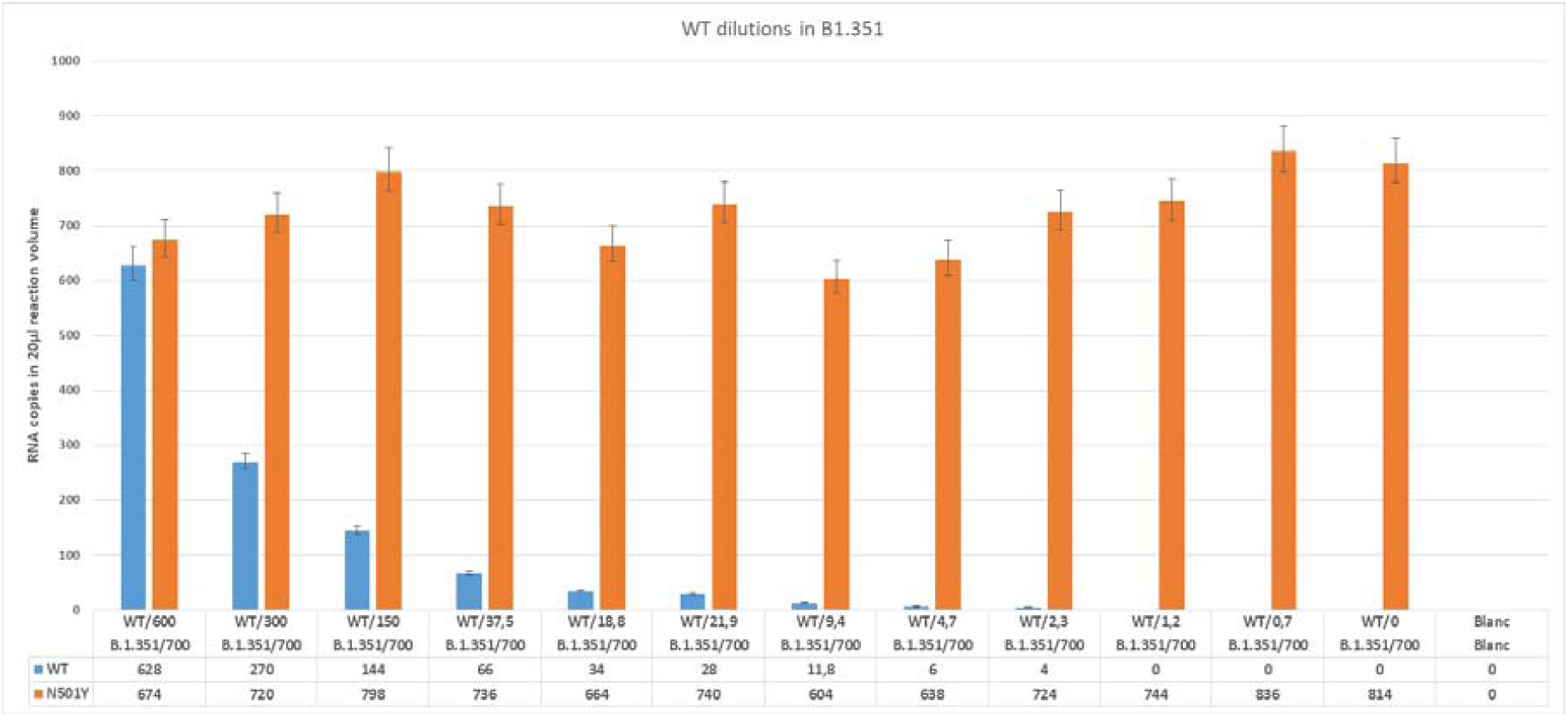
Detected copy numbers of WT and N501Y in samples containing ~700 copies variant lineage B.1.351WT and 2-fold dilution series of WT.

These results demonstrate the ability of the method to simultaneously detect and discriminate between the sequences of WT and the N501Y mutation in lineage B.1.351 from a mixed sample. It also shows that low concentrations of N501Y mutation can be detected in the presence of WT virus RNA and vice versa. Detection of N501Y was possible in a sample dilution containing a theoretical concentration of only 2.7 copies of B.1.351 or WT detection in a sample containing 2.3 copies of WT suggests the feasibility for specific detection of low concentrations of WT and N501Y.

Comparing the measured versus expected proportion of the 2-fold dilutions of B.1.351 (Figure 3 A) or the diluted WT (Figure 3B) shows the high linear correlation between the expected proportion and detected proportion of N501Y and WT respectively.

**Figure 3.**
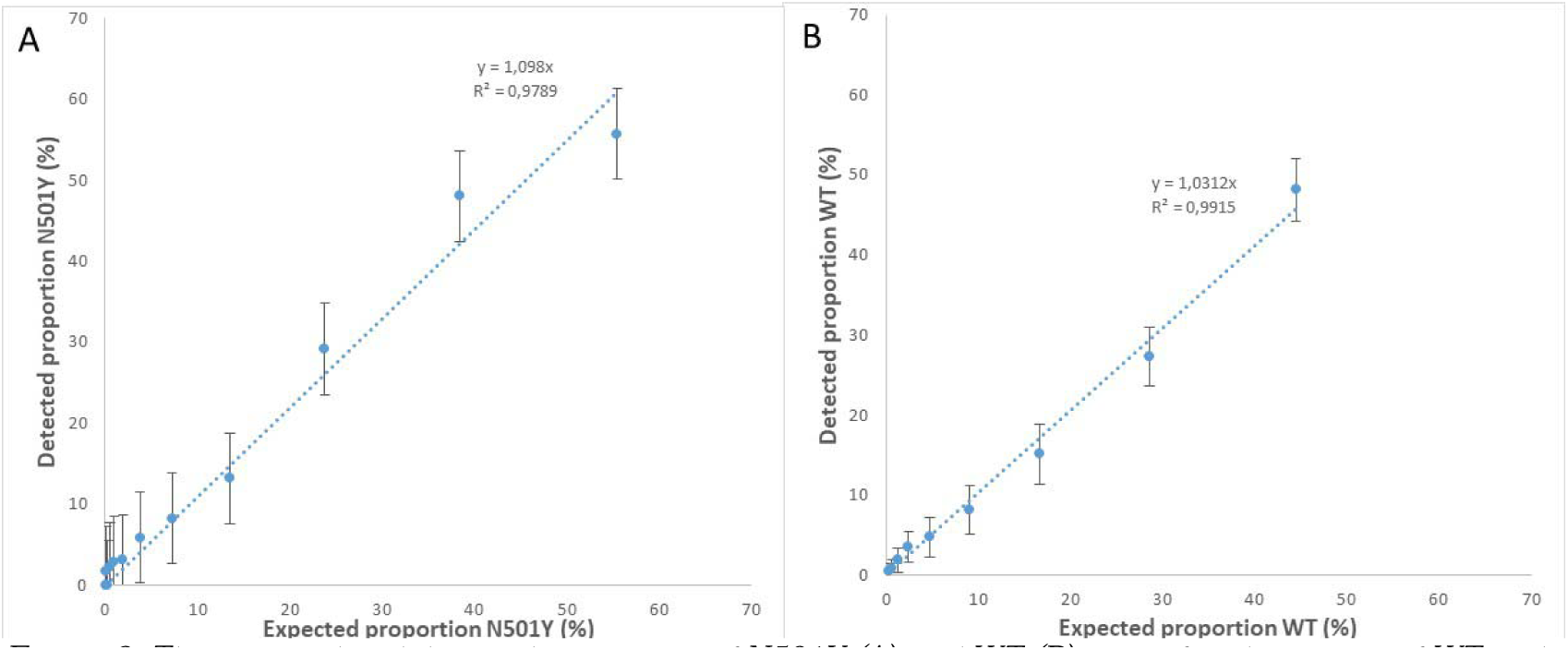
The expected and detected proportion of N501Y(A) and WT (B) in artificial mixtures of WT and lineage B.1.351 as detected by ddPCR.

These results demonstrate that the assay can be used to discriminate between WT SARS-CoV-2 RNA and variants containing the N501Y mutation and simultaneous detection and quantification of these sequences. The specificity of the assay to detect the N501Y mutation was confirmed on RNA from SARS-CoV-2 lineage B.1.1.7 (data not shown). It is expected that the assay can be used to detect low concentrations of WT and N501Y and to obtain reliable insight in the proportion of N501Y in mixtures of viruses, and therefore has the potency to collect information about the spread of variants.

### SARS-CoV-2 VoC surveillance in wastewater of Amsterdam and Utrecht

Wastewater samples from the cities of Amsterdam and Utrecht from September 2020 to March 2021 were analyzed to evaluate the applicability of RT-ddPCR to monitor the emergence of VoC’s via wastewater. The concentration SARS-CoV-2 N2 RNA in these samples was first determined using RT-qPCR and compared to newly reported COVID-19 cases in these cities.

#### SARS-CoV-2 sewage surveillance in Amsterdam and Utrecht

The concentrations of SARS-CoV-2 N2 (normalized for the concentration of CrAssphage in the same sample) in Amsterdam and Utrecht wastewater and the incidence of newly reported cases in Amsterdam and Utrecht are depicted in Figure 4. The results show that the concentrations of SARS-CoV-2 RNA in wastewater reflected the trends in the newly reported positive COVID-19 clinical tests in the population of these cities. Both cities experienced a peak in COVID-19 cases in October and December 2020.

**Figure 4.**
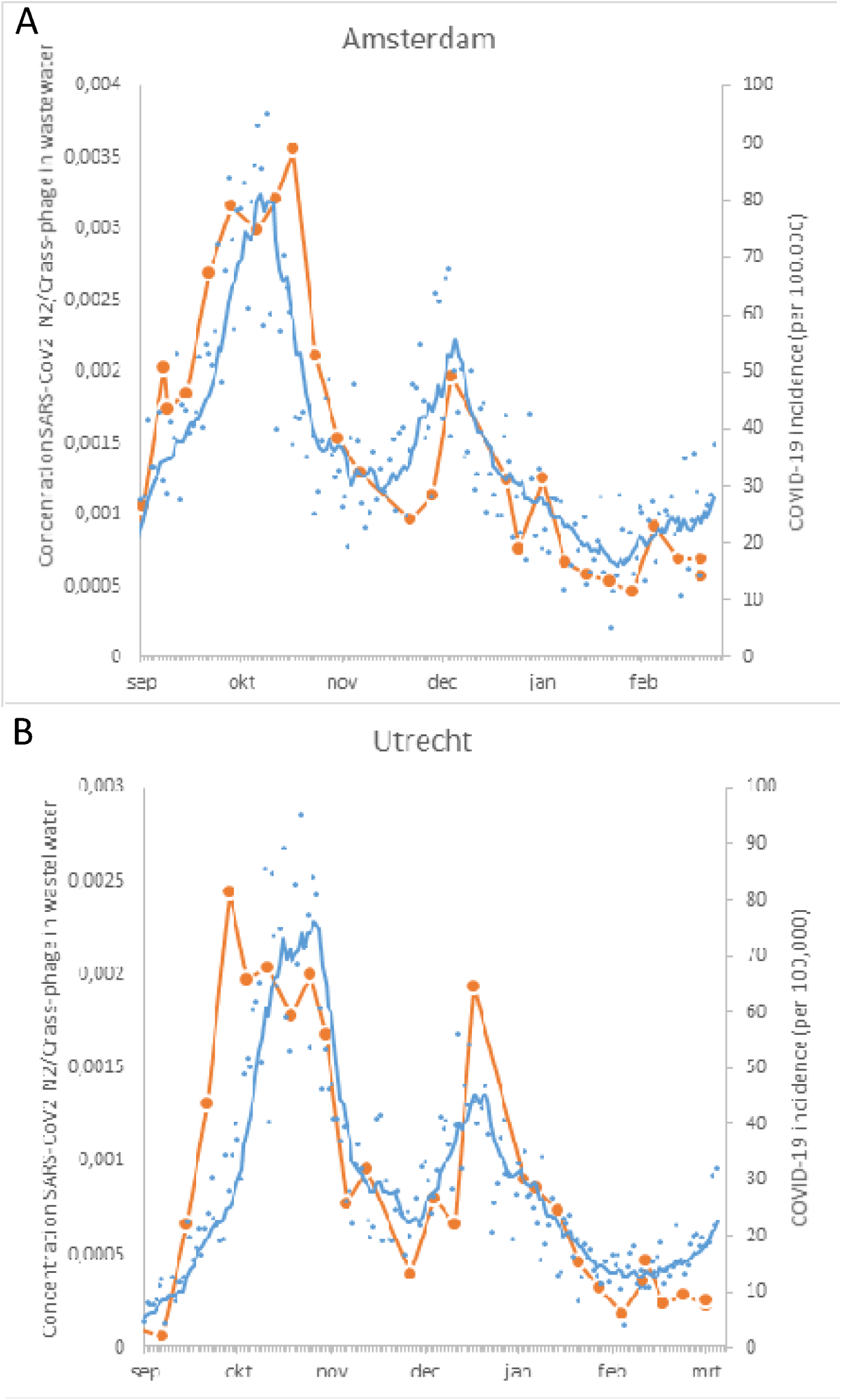
The concentration of SARS-CoV-2 N2 gene (normalized for the concentration of Crass-phage) in wastewater of Amsterdam (A) and Utrecht (B) from Aug 26 2020 to March 8 2021 (orange circles) and the newly reported COVID-19 cases in the city population (blue dots) and the 7-day moving average (blue line) COVID-19 data: National Institute of Public Health and the Environment, data source ESRI NL COVID-19 Hub (Esri-Nederland 2021)

#### Emergence of the SARS-CoV-2 N501Ymutation in sewage from Amsterdam and Utrecht

Analysis of the sewage samples was performed to study the applicability of the N501Y RT-ddPCR assay. The results are summarized in table S1 (Amsterdam) and table S2 (Utrecht). The proportion of N501Y to WT sequences in Amsterdam and Utrecht wastewater was compared with publicly available data from the Dutch National Institute for Public Health and the Environment (RIVM). The available data is based on surveillance of variants by sequencing a random selection of SARS-COV-2 isolates from several hundreds of new cases in the Netherlands every week (Table S3, “Pathogen Genomic surveillance”: https://www.rivm.nl/coronavirus-covid-19/virus/varianten, accessed March 09 2021). The results of these comparisons are shown in Figures 5A (Amsterdam) and 5B (Utrecht).

**Figure 5.**
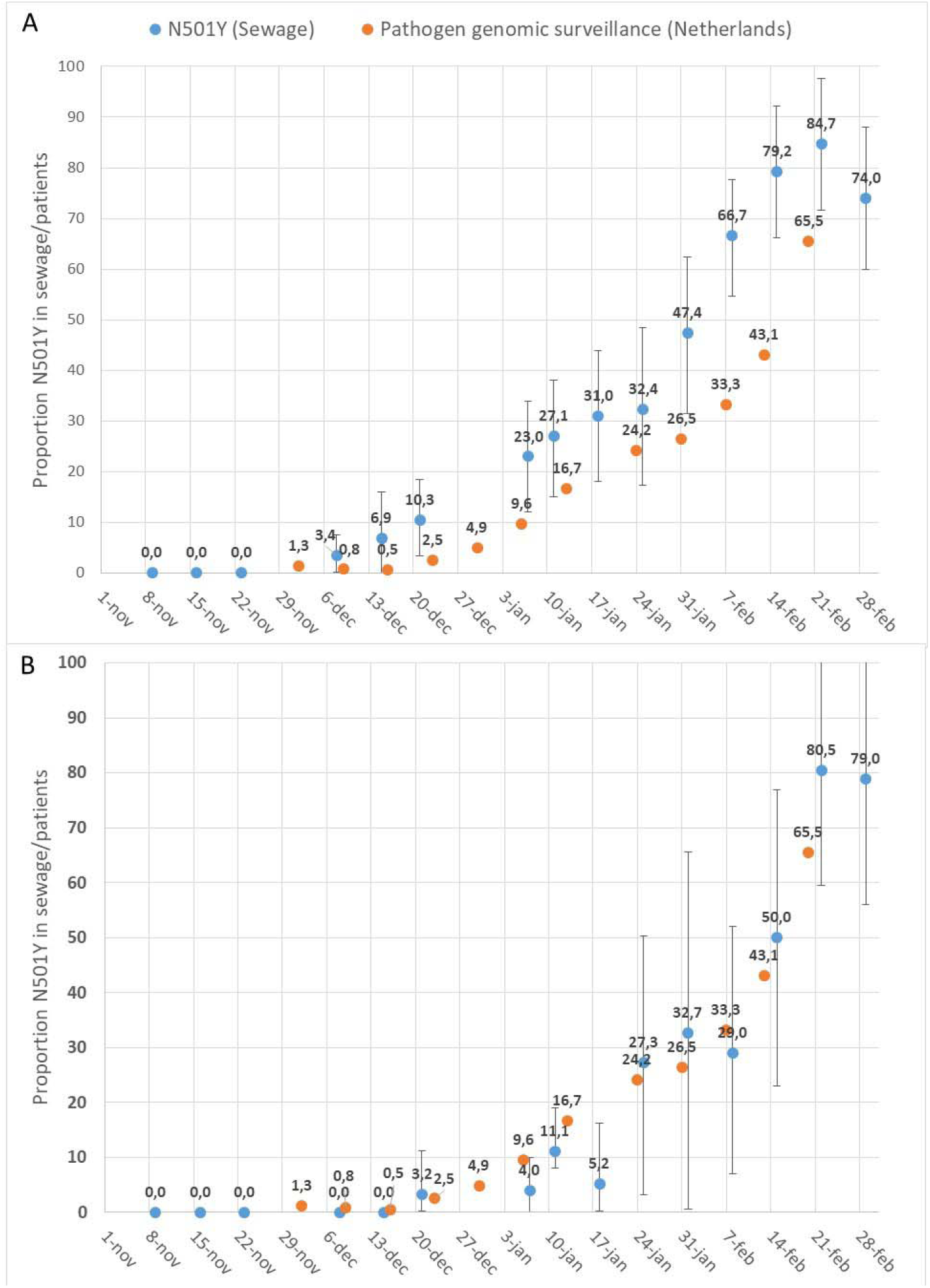
Proportion of Spike gene RNA fragments containing the N501Y mutation in wastewater from Amsterdam (A) and Utrecht (B) calculated as relative concentration of N501Ycontaining Spike gene fragments divided by the total concentration of S-gene fragments (WT+ N501Y). Error bars are calculated assuming Poisson distribution of RNA molecules in droplets. The proportion of newly COVID-19 patients infected with N501Y containing variants (Pathogen genomic surveillance) are shown as (the proportional sum of lineages B.1.1.7, B.1.351 and P.1).

The earliest detection of N501Y was in the wastewater sample of December 8 2020 in Amsterdam and in the wastewater sample of December 21 2020 in Utrecht. The proportion of N501Y gradually increased to 85% on February 22 2021 in Amsterdam and 81% on February 22 2021 in Utrecht. The total SARS-CoV-2 RNA concentrations were higher in wastewater from Amsterdam compared to Utrecht (Table S1 and S2), yielding a more accurate assessment of the N501Y proportion, as indicated by the error-bars.

The first detection of the British variant (lineage B.1.1.7) in patients in the Netherlands was in the first week of December 2020. This corresponds to the first detection of N501Y in wastewater from Amsterdam. The first detection of N501Y in wastewater from Utrecht was two weeks later. After the emergence of variants containing the N501Y mutation, the increase in the proportion of this marker in wastewater follows a trend that is comparable with the increase of N501Y containing variants in patients in the Netherlands. The percentage of N501Y containing variants appears to be higher in Amsterdam wastewater in the complete period suggesting earlier introduction of the British variant in Amsterdam than in Utrecht. The introduction and emergence of SARS-CoV-2 variants containing the N501Y mutation in Utrecht wastewater aligns more in time with the national ‘average’ emergence as obtained through the genomic pathogen surveillance data.

#### Comparison RT-qPCR with RT-ddPCR

The concentrations SARS-CoV-2 RNA measured in sewage from WWTP Amsterdam and Utrecht with RT-qPCR (N2 amplicon of the N-gene) and RT-ddPCR (sum of WT and N501Y sequences of the Spike gene) are compared in Figure 6. This comparison shows that there is a linear relation between the values measured with both methods. However, concentrations measured with RT-qPCR are consistently higher (61% on average) than concentrations measured with RT-ddPCR. On basis of current knowledge of SARS-CoV-2 sequences in the Dutch population, it is unlikely that there are other sequence variants in the populations that were detected by RT-qPCR but missed with the RT-ddPCR assay. The higher RT-qPCR results could be the result of targeting different gene fragments with potential differences in detected concentrations in wastewater. The different RNA-targets can have different degradation rates in sewage but it can also be the result of higher expression levels of the N-gene compared to the S-gene in coronavirus infected cells and the presence of the N-gene in every subgenomic mRNA (Kim, Lee et al. 2020) making it likely that higher levels of N-sequences will be present in infected intestinal cells and eventually may lead to higher N-sequence concentrations in sewage.

**Figure 6.**
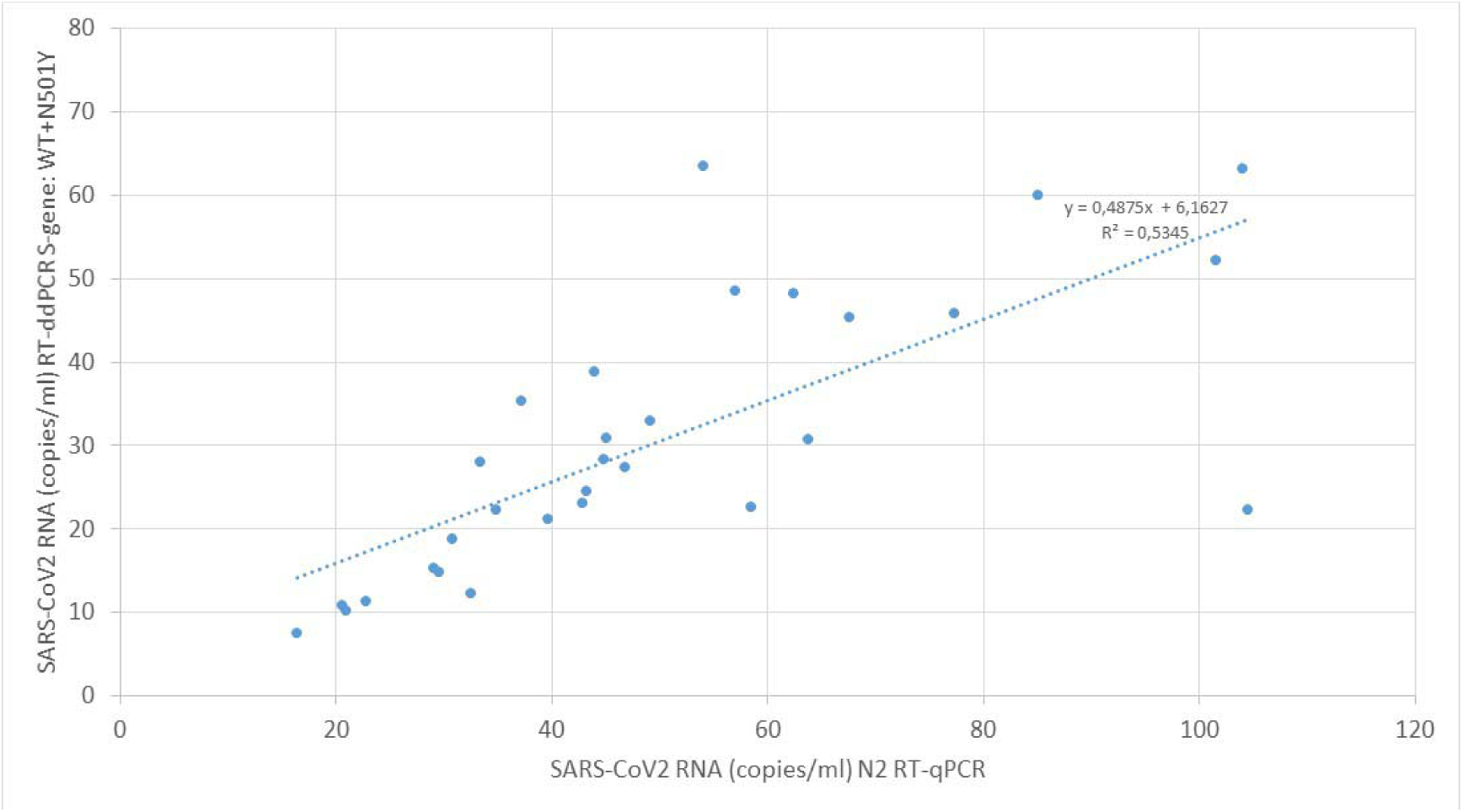
Comparison of SARS-CoV-2 RNA, measured with RT-qPCR and RT-ddPCR, concentrations in sewage.

## Discussion and conclusions

This research describes the first use of RT-ddPCR to specifically detect and quantify SARS-CoV-2 mutations associated with VoC’s in wastewater. This assay can be used for sensitive and simultaneous detection of WT and the N501Y mutation in sewage. Although more extensive research is needed to determine the limit of detection accurately, experiments on SARS-CoV-2 RNA from lineage B.1.351 (with mutation N501Y) and WT in different proportional mixtures demonstrated that proportions as low as 0,5% of lineage B.1.351 can be detected in a background of 700 RNA-copies of WT SARS-CoV-2. These experiments also demonstrated the linearity of the expected proportion and the measured proportion of B.1.351 in the mixtures and vice versa, indicating that this N501Y assay can be used to accurately determine the ratio between SARS-CoV-2 genomes containing N501Y and those containing the WT sequence. The weekly analysis of wastewater from the cities of Amsterdam and Utrecht using RT-qPCR targeting the N-gene (CDC N2 target) clearly demonstrated the relationship between SARS-CoV-2 RNA levels and the Covid-19 incidence in the communities connected to the respective WWTP’s. N501Y containing variants were first detected on December 8 2020 in Amsterdam wastewater and two weeks later in Utrecht wastewater. The proportion of N501Y variants in the weekly samples increased to approximately 80% on March 1, 2021 in both cities.

These results demonstrate the applicability of this RT-ddPCR assay to monitor the absolute quantities and ratios of WT viral sequences and sequences containing the N501Y mutation. This makes the use of RT-ddPCR on wastewater samples an attractive, rapid and efficient method to follow the emergence of (mutations associated with) VoC’s in the community, that can generate an early warning of the emergence of VoC’s on the basis of one “population-sample” via domestic wastewater. This proof-of-principle demonstrates the value of RT-ddPCR to detect N501Y, and suggests RT-ddPCR can also be used for the detection of multiple mutations (like f.e. E484K, K417 or P681H) characteristic for various epidemiologically or immunologically relevant VoC’s in sewage to monitor the emergence and spread of VoC in the community. Whole genome sequencing of SARS-CoV-2 from clinical samples from infected individuals is commonly used to identify VoC’s (Tegally, Wilkinson et al. 2021). Although whole genome sequencing has shown to be feasible to give insight in the sequence variations of SARS-CoV-2 genomes in sewage (Izquierdo-Lara, Elsinga et al. 2020) and has been used to detect mutations associated with variants of concern in wastewater(Jahn, Dreifuss et al. 2021), this requires deep sequencing and bioinformatics to determine the presence of mutations associated with VoC’s in wastewater, and is provides less quantitative results than ddPCR. An advantage of whole genome sequencing of wastewater samples is that it can provide more extensive information about the co-occurrence of the range of mutations and deletions associated with VoC. It would be valuable to compare the results of RT-ddPCR with the N501Y (and other) mutations with those of whole genome sequencing on the same wastewater samples and on samples of the COVID-19 cases in the population that contributes to this wastewater.

## Data Availability

All data details described in this paper is available and can be provided upon request

## Acknowledgements

The authors are very grateful for the assistance of the Water Authorities and WWTP operators of Waternet and Hoogheemraadschap de Stichtse Rijnlanden in the Netherlands who organized the sampling and provided the sewage samples. We are also grateful to Meindert de Graaf for sample transport. Eddy van Collenburg (BIORAD) is acknowledged for technical advice and providing the test kits. Coronavirus MHV-A59 was a kind gift from Prof. Eric Snijder and provided by Linda Boomaars (Leids Universitair Medisch Centrum). This study was financed by TKI Health Holland in EUREKA project WASTEWATER4COVID.

## Supplemental material

**Table S1.** Summary of the RT-ddPCR N501Y and WT results in wastewater samples from WWTP Amsterdam

| Amsterdam<br>Date | RT-qPCR |  |  | RT-ddPCR |  |  |  |  |  |  |  |
| --- | --- | --- | --- | --- | --- | --- | --- | --- | --- | --- | --- |
|  | Analyzed<br>sewage volume<br>(ml) | Recovery<br>MHV-RNA (%) | SARS-CoV2 (N2)<br>conc. (copies/ml) | Droplets<br>in reaction | RNA-copies in 20 µl RT-ddPCR reaction |  |  | Concentration in Sewage (RNA-copies/ml) |  |  | Fractional<br>abundance (%)<br>N501Y |
|  |  |  |  |  | N501Y | WT | N501Y+WT | N501Y | WT | N501Y+WT |  |
| 9-11-2020 | 2,8 | 25,2 | 85,1 | 14540 | 0 | 162 | 162 | 0,0 | 60,0 | 60,0 | 0,0 |
| 16-11-2020 | 2,7 | 29,0 | 101,5 | 15274 | 0 | 142 | 142 | 0,0 | 52,2 | 52,2 | 0,0 |
| 23-11-2020 | 2,7 | 18,6 | 57,0 | 10425 | 0 | 130 | 130 | 0,0 | 48,6 | 48,6 | 0,0 |
| 8-12-2020 | 2,7 | 10,1 | 54,0 | 11790 | 6 | 168 | 174 | 2,2 | 61,3 | 63,5 | 3,4 |
| 15-12-2020 | 2,7 | 10,4 | 39,7 | 12018 | 4 | 54 | 58 | 1,5 | 19,6 | 21,1 | 6,9 |
| 21-12-2020 | 2,7 | 13,3 | 104,0 | 9183 | 18 | 156 | 174 | 6,5 | 56,7 | 63,2 | 10,3 |
| 7-1-2021 | 2,8 | 26,3 | 67,6 | 10636 | 28 | 94 | 122 | 10,4 | 34,9 | 45,4 | 23,0 |
| 11-1-2021 | 2,7 | 10,7 | 37,2 | 13842 | 26 | 70 | 96 | 9,6 | 25,7 | 35,3 | 27,1 |
| 18-1-2021 | 2,7 | 19,8 | 63,7 | 13278 | 26 | 58 | 84 | 9,5 | 21,2 | 30,7 | 31,0 |
| 25-1-2021 | 2,7 | 14,0 | 43,3 | 12228 | 22 | 46 | 68 | 7,9 | 16,6 | 24,5 | 32,4 |
| 1-2-2021 | 2,8 | 12,9 | 33,4 | 12768 | 36 | 40 | 76 | 13,3 | 14,7 | 28,0 | 47,4 |
| 8-2-2021 | 2,7 | 27,8 | 62,5 | 12252 | 88 | 44 | 132 | 32,1 | 16,0 | 48,1 | 66,7 |
| 15-2-2021 | 2,7 | 14,5 | 34,9 | 14939 | 48 | 13 | 61 | 17,6 | 4,6 | 22,3 | 79,2 |
| 22-2-2021 | 2,7 | 19,3 | 58,5 | 12449 | 52 | 9 | 61 | 19,2 | 3,5 | 22,6 | 84,7 |
| 1-3-2021 | 2,7 | 7,6 | 44,8 | 12284 | 56 | 20 | 76 | 20,9 | 7,5 | 28,4 | 74,0 |
|  |  |  | Pos control WT | 18242 | 0 | 652 | 652 |  |  |  | 0,0 |
|  |  |  | Pos control N501Y | 17088 | 668 | 0 | 668 |  |  |  | 100,0 |
|  |  |  | Blank sample | 16161 | 0 | 0 | 0 |  |  |  | 0,0 |

**Table S2.**
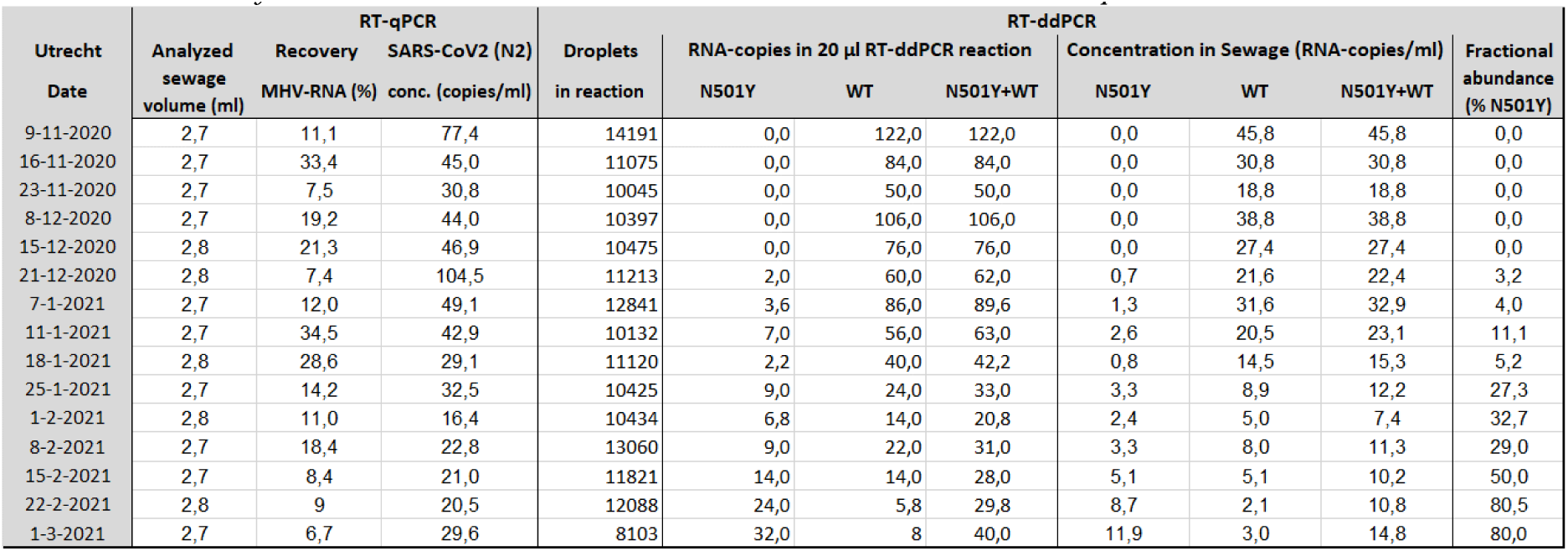
Summary of the RT-ddPCR N501Y and WT results in wastewater samples from WWTP Utrecht

**Table S3.** N501Y containing SARS-CoV-2 isolates based on sequence analysis of isolates from patients (“ Pathogen Genomic surveillance”: https://www.rivm.nl/coronavirus-covid-19/virus/varianten, accessed March 09 2021)

| Week number | Number of sequenced samples | British variant* | South African variant* | Brasilian variant P1* | British variant containing E484K mutation* | Variant B.1.525 + E484K + F888L | Variants containing N501Y | % containing N501Y |
| --- | --- | --- | --- | --- | --- | --- | --- | --- |
| 2020/49 | 77 | 1 |  |  |  |  | 1 | 1,3 |
| 2020/50 | 118 | 1 |  |  |  |  | 1 | 0,8 |
| 2020/51 | 190 | 1 |  |  |  |  | 1 | 0,5 |
| 2020/52 | 238 | 5 | 1 |  |  |  | 6 | 2,5 |
| 2020/53 | 204 | 9 | 1 |  |  |  | 10 | 4,9 |
| 2021/1 | 334 | 32 |  |  |  |  | 32 | 9,6 |
| 2021/2 | 270 | 43 | 2 |  |  |  | 45 | 16,7 |
| 2021/3 | 576 | 131 | 3 |  |  |  | 134 | 23,3 |
| 2021/4 | 786 | 194 | 14 | 1 |  |  | 209 | 26,6 |
| 2021/5 | 812 | 252 | 17 |  | 1 |  | 270 | 33,3 |
| 2021/6 | 881 | 355 | 26 |  |  |  | 381 | 43,2 |
| 2021/7 | 811 | 497 | 16 |  | 1 | 4 | 518 | 63,9 |
| 2021/8 | 588 | 442 | 11 | 1 |  | 2 | 456 | 77,6 |
| * Variant containing N501Y |  |  |  |  |  |  |  |  |

